# Determinants of maternal morbidity during pregnancy in urban Bangladesh: Negative binomial regression approach

**DOI:** 10.1101/2022.05.03.22274640

**Authors:** Zakir Hossain, Nilima Afroz, Sabina Sharmin, Sayema Sharmin, Enamul Kabir

**Affiliations:** Department of Statistics, University of Dhaka, Dhaka, Bangladesh; Road Transport and Highways Division, Ministry of Road Transport and Bridges, Dhaka, Bangladesh; School of Mathematics, Physics and Computing, University of Southern Queensland, Toowoomba, Australia

**Keywords:** Count Data, Overdispersion, Mixed Models, Incidence Rate Ratio (IRR)

## Abstract

**Aim:** To investigate the prevalence of maternal morbidity during pregnancy and its determinants among the women from urban areas of Bangladesh.

**Methods:** The secondary data were used and extracted from the latest Bangladesh Urban Health Survey (BUHS) 2013. Several statistical models: Poisson, negative binomial (NB) and mixed Poisson were adapted and compared to explore the best model for investigating potential determinants of maternal morbidity. Pearson chi-square statistic was used for the detection of overdispersion in the data.

**Results:** Overall 13.5% of the urban women in Bangladesh suffered from at least two pregnancy complications. The study detected the overdispersion existing in the maternal morbidity count data and found the NB regression as the best choice for analyzing the data because of its smallest Akaike information criterion. Administrative division (Rangpur: *p*=0.003, IRR=1.34, 95% CI: 1.11 to 1.63; Sylhet: *p*=0.006, IRR=1.42, 95% CI: 1.11 to 1.82), wanted pregnancy (p<0.001, IRR=0.80, 95% CI: 0.71 to 0.90), place of delivery (*p*<0.001, IRR=0.60, 95% CI: 0.54 to 0.66) and wealth index (Rich: *p*<0.001, IRR=0.74, 95% CI: 0.66 to 0.84) were found to be statistically significant determinants for maternal morbidity during pregnancy among the urban women in Bangladesh.

**Conclusions:** The urban women in Bangladesh with unwanted pregnancy, from poor/middle income group; and living in Rangpur and Sylhet divisional cities have higher risk of maternal morbidity during pregnancy. The women already suffering from major pregnancy related complications visit health centre to give birth. Study findings may help the government and relevant authorities to take necessary steps for reducing maternal morbidity and mortality due to pregnancy related complications.

## Introduction

Reproductive health care is a highly focused issue in the development of a country. Millions of women experience life-threating and other health related complications during pregnancy, delivery and post-partum periods in developing countries like Bangladesh. Despite pregnancy related complications are commonly seen among women in Bangladesh, not too many attentions have been drawn by the researchers and other relevant authorities to reduce such complications, and hence maternal and child mortality. Like other developing countries, a rapid urbanization problem in Bangladesh is growing in recent decades as a part of the universal trend. In 2015, about 65.7% of the total population was living in rural areas however, by 2039 the majority of population will live in urban areas of Bangladesh [1].

Approximately 3.8% of total population in Bangladesh or above 5.7 million people are living in the urban slums with very limited health facilities [2]. Although health care services are available in most urban regions of Bangladesh, the underprivileged residents in some urban parts have fewer access to important health care services than those from wealthier areas. The community health centre facility was freely available only for 7.3% of slum dwellers in Bangladesh [3]. In urban slum areas, the poorest and illiterate women were less likely to take adequate maternal health care services from the medically skilled professionals [4].

Every year, about 9.5 million women are suffering from pregnancy and delivery related complications worldwide and the number of maternal deaths is estimated to be more than 300,000 [5, 6]. In 2015, the maternal mortality rate (MMR) of Bangladesh was 176 while the targeted MMR for the millennium development goals (MDGs) was set to reduce at 143 per 100,000 live births, and this rate was substantially higher in Bangladesh compared to other countries [7]. Moreover, the global MMR is fixed in the 2030 sustainable development goal (SDG 3.1) to less than 70 per 100,000 live births [8]. Thus, investigating the reasons of maternal mortality in Bangladesh is very important for further reduction in MMR targeting its SDG 3.1.

Approximately three quarters of maternal mortality are occurred due to pregnancy and delivery related complications [9]. About 810 women died every day in 2017 from avoidable causes of pregnancy and childbirth related complications and approximately 94% of these maternal deaths happen in low and lower middle-income countries [8]. A study on maternal mortality in Bangladesh reported that women education and poverty were two significant factors for maternal deaths [10]. Another recent study also found that early maternal age (<18 years), unwanted pregnancy, migration status and NGO (non-government organization) membership were potential risk factors for pregnancy and delivery related complications among the urban women in Bangladesh [11].

Hemorrhage/severe bleeding, fits/convulsion, odema, excessive vomiting, and cough/high fever were considered as life-threatening and high risk pregnancy or delivery complications [12–15]. Thus, it is highly important to investigate associated factors for these complications to reduce maternal and child mortality and hence to give safe births to achieving the SDGs. Unfortunately, a limited number of studies has been conducted directly addressing the maternal morbidity/complications during pregnancy in the urban areas of Bangladesh.

The main objective was to find out the potential determinants of maternal morbidity during pregnancy among the urban women in Bangladesh using the latest Bangladesh Urban Health Survey (BUHS) 2013 data. The number of pregnancy complications was considered as the count response variable and therefore, overdispersion (extra-variation) nature of the data has also been explored, entirely overlooked in previous studies, and taken into account in the current study to avoid misleading inferences and hence for valid interpretation of the results.

## Materials and methods

### Data and sampling design

We used the secondary data on maternal morbidity during pregnancy, extracted from the latest Bangladesh Urban Health Survey (BUHS) 2013. The survey was conducted using a three-stage stratified sampling design during 23 July to 12 December, 2013 and the details are available at https://dataverse.unc.edu/. The data were collected from 1718 clusters in major urban areas, comprising of 450 in City-corporation slums, 900 in City-corporation non-slums and 368 in other urban areas of Bangladesh. In total, after excluding missing cases, 6001 women records were used in this study who had their last birth preceding three years of the BUHS 2013.

### Variables included in the Study

We considered different types of pregnancy morbidities or complications, namely, haemorrhage/severe bleeding, fits/convulsion, odema, prolonged labour, high fever, leaking membrane or fluid, mal presentation, retained placenta, high blood pressure and severe headache experienced by the urban women during their last pregnancy of child birth [11, 15]. The count response variable of pregnancy complications is our interest in this study.

To investigate the potential risk factors of maternal morbidity during pregnancy, several socio-economic and demographic variables: age, administrative division or place of residence, religion, education, ever-born children, multiple last birth, wanted pregnancy, at least 8 ANC (antenatal visits) [16, 17], place of delivery, delivery by MTP (medically trained professional), sex of last child, migration status, wealth index, NGO (non-government organization) membership and media exposure are considered as the explanatory variables. The women came from rural areas or other cities were considered as migrant. The variables ‘media exposure’ and ‘NGO membership’ were derived by combining the associated covariates available in the survey as these were not found directly from the survey data. Women who read a newspaper or magazine or listen to radio or watch television were considered as exposed to media. Those women who were allied with any one of the organizations: Grameen Bank, Bangladesh Rural Advancement Committee (BRAC), Bangladesh Rural Development Board (BRDB), Association of Social Advancement (ASA) and Proshika were considered as NGO members.

### Overdispersion

Overdispersion is a very common scenario in modelling count data. It occurs for the greater variability i.e. when the variance of responses in a Poisson regression model is higher than the mean. Overdispersion should be taken into account for analyzing pregnancy complications count data of women used in this study to avoid misleading inferences. Overdispersion can be detected by using the value of Pearson chi-square (χ^2^) statistic divided by the associated degrees of freedom (df) [18]. This value is called as the *dispersion* and used as 1, >1 and <1 for equidispersed, overdispersed and underdispersed models, respectively. The Pearson χ^2^ statistic can be written as

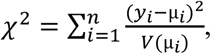

where *y*_*i*_ and µ_*i*_ denote observed and expected counts, respectively; *V* is known as the variance function which is equal to µ_*i*_ for Poisson and µ_*i*_ + *γ*µ_*i*_^2^ for the negative binomial regression models with the dispersion parameter *γ* [18].

### Models for count data

We considered several count regression models for analyzing the maternal morbidity data of women during their pregnancy in urban Bangladesh. More precisely, Poisson regression (PR), negative binomial regression (NBR) and mixed Poisson regression (MPR) models were adapted in the context of generalized liner models (GLM) and generalized liner mixed models (GLMM) framework. The PR is the initial step for modelling count data. Let *Y*_*i*_; (*i* = 1, …, *n*) be the count response for *i*^th^ individual with mean *E*(*Y*_*i*_) = µ_*i*_, then the PR model using the *log* link-function can be written in the GLM framework [19, 20] as

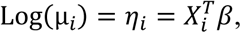

where *X*_*i*_ denotes a *p* × 1 column vector of covariates, *β* is a 1× *p* vector of regression parameters and *η*_*i*_ is called the linear predictor. The PR model is restrictive because of its equidispersion assumption of the mean and variance of count responses. However, count data are often overdispersed (extra variation) or underdispersed (less variation) compared to the PR model.

The negative binomial regression (NBR) is an alternative and widely used to model over-dispersed count data. The random effect *U* is considered to model the unobserved variability exists in the data. It is usually assumed that *U* follows the gamma distribution mainly for computational simplicity with shape parameter *δ* and scale parameter *δ* ^−1^. The variance of *Y*_*i*_ in NBR model can be written as *Var*(*Y*_*i*_) = µ_*i*_ + *γ*µ_*i*_^2^ = µ_*i*_(1 + *γ*µ_*i*_) with *γ* >0 [18, 21]. The overdispersion i.e. the extra quantity of µ_*i*_ is 1 + *γ*µ_*i*_, multiplicative factor, which depends on µ_*i*_. It shows that the variance is larger than the mean and therefore, the overdispersion in the data is accounted for the NBR model in contrast to the PR through the dispersion parameter *γ* = *δ* ^−1^. When *γ* = 0, then *Var*(*Y*_*i*_) = µ_*i*_ and hence *E*(*Y*_*i*_) =*Var*(*Y*_*i*_) = µ_*i*_, which is the condition of equidispersed Poisson distribution.

The MPR is also a further improvement of modelling correlated or clustered count data, which is an extension of the PR where random effects are introduced in the linear predictor [22]. Let *Y*_*jk*_ be the *k*^th^ individual (*k* = 1, …, *n*_*j*_) under j^th^ cluster (*j* = 1, …, *m*) then similar to the PR, the MPR can be written in the context of GLMM as

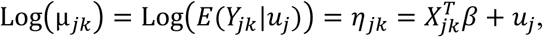

where µ_*jk*_ = *E*(*Y*_*jk*_|*u*_*j*_) is the conditional expectation and *η*_*jk*_ is the liner predictor [23, 24]. As before *log* is the link function and *u*_*j*_ is the *j*^th^ random effect. Estimates of the model parameters are obtained by assuming normally distributed and uncorrelated random effects. The likelihood-based model selection criterion, Akaike information criterion (AIC), is used to select the best model with minimum AIC value and it can be defined as

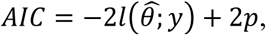

where 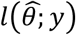 is the log-likelihood, 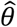 is the vector of estimated regression parameters and *p* is the number of model parameters of interest[25]. For the convenience of interpretation of estimated model parameters, the incidence rate ratio (IRR) associated with the individual covariate *X*_*i*_; *i* = 1, …, *p* is widely used and can be written as 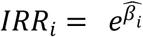, where 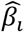 is the *i*^th^ estimated regression coefficient [26].

### Ethics statement

This article does not contain any studies with human participants performed by the author. The Bangladesh Urban Health Survey (BUHS) 2013 was approved by ICF Macro Institutional Review Board and the National Research Ethics Committee of the Bangladesh Medical Research Council. A written consent about the survey was given to participants before conducting the interview. All identification of the respondents was disidentified before publishing data. The secondary data set was used in this study and freely available online at https://dataverse.unc.edu/.

## Results

Table 1 shows that among all urban women included in this study, 3.3% (197 out of 6001) and 2.7% (161 out of 6001) experienced life-threating pregnancy complications, hemorrhage/severe bleeding and fits/convulsion, respectively. It can also be seen that 15.3% (917 out of 6001), 7.6% (455 out of 6001) and 1.2% (73 out of 6001) of Bangladeshi urban women faced high-risk pregnancy complications: odema, prolonged labor and high fever, respectively.

**Table 1:** Frequency and percentage (%) distributions of pregnancy complications among the urban women (n=6001) in Bangladesh

| Pregnancy complications | % |
| --- | --- |
| Hemorrhage Severe bleeding | 3.3 |
| Fits/Convulsion | 2.7 |
| Odema | 15.3 |
| Prolonged labor | 7.6 |
| High fever | 1.2 |
| Leaking membrane | 6.7 |
| Mal presentation | 1.1 |
| Retained placenta | 1.2 |
| High blood pressure | 5.5 |
| Severe headache | 12.4 |

In addition, we can observe that 6.7% (401 out of 6001) urban women faced the complexity of leaking membrane, 1.1% (67 out of 6001) mal-presentation, 1.2% (69 out of 6001) retained placenta, 5.5% (331 out of 6001) high blood pressure and 12.4% (747 out of 6001) severe headache or blurred vision. The number of pregnancy complications/morbidities with its mean and variance were computed and summarized in Table 2.

**Table 2:** Distribution of the number of pregnancy complications among the urban women in Bangladesh

| Distribution | Number of pregnancy morbidities/complications |  |  |  |  |  | Total |
| --- | --- | --- | --- | --- | --- | --- | --- |
|  | 0 | 1 | 2 | 3 | 4 | 5+ |  |
| Frequency | 3851 | 1339 | 515 | 185 | 72 | 39 | 6001 |
| % | 64.2 | 22.3 | 8.6 | 3.1 | 1.2 | 0.6 | 100 |

Mean=0.57 and variance=0.90 (i.e. extra variation)

Table 2 reveals that 22.3% (1339 out of 6001) of the urban women in Bangladesh experienced one and 13.5% at least two complications during their pregnancy. Bivariate analysis was carried out considering several socio-economic, demographic and biological variables associated with pregnancy complications using the ANOVA (analysis of variance) F-test (Table 3). Administrative division (*p*<0.001), education (*p*=0.032), wanted pregnancy (*p<*0.001), place of delivery (*p<*0.001), wealth index (*p<*0.001) and NGO membership (*p<*0.001) of women were found to be statistically significant at 5% level with their mean number of pregnancy complications.

**Table 3:** Bivariate analysis of the socio-economic and demographic variables along with 95% confidence interval (CI) for mean number of pregnancy complications along with *p*-values of the ANOVA *F*-tests.

| Variable | Mean morbidity | 95% CI | $p$-value |
| --- | --- | --- | --- |
| Age (years) |  |  | 0.505 |
| <=20 | 0.59 | (0.53, 0.64) |  |
| >20 | 0.57 | (0.54, 0.59) |  |
| Administrative division |  |  | **<0.001** |
| Dhaka | 0.57 | (0.53, 0.60) |  |
| Barisal | 0.72 | (0.56, 0.88) |  |
| Chittagong | 0.49 | (0.44, 0.53) |  |
| Khulna | 0.62 | (0.52, 0.72) |  |
| Rajshahi | 0.56 | (0.48, 0.65) |  |
| Rangpur | 0.80 | (0.68, 0.92) |  |
| Sylhet | 0.75 | (0.57, 0.94) |  |
| Religion |  |  | 0.520 |
| Non-Muslim | 0.54 | (0.46, 0.63) |  |
| Muslim | 0.57 | (0.55, 0.60) |  |
| Education |  |  | **0.032** |
| Primary | 0.61 | (0.56, 0.65) |  |
| Secondary | 0.57 | (0.54, 0.60) |  |
| Higher | 0.51 | (0.47, 0.56) |  |
| Ever born children |  |  | 0.979 |
| 1 or 2 | 0.57 | (0.54, 0.60) |  |
| 3 or higher | 0.57 | (0.51, 0.63) |  |
| Multiple last birth |  |  | 0.438 |
| No | 0.70 | (0.30, 1.10) |  |
| Yes | 0.57 | (0.54, 0.59) |  |
| Wanted pregnancy |  |  | **<0.001** |
| No | 0.71 | (0.64, 0.79) |  |
| Yes | 0.55 | (0.52, 0.57) |  |
| At least 8 ANC visits |  |  | 0.160 |
| No | 0.57 | (0.54, 0.59) |  |
| Yes | 0.65 | (0.53, 0.76) |  |
| Media exposure |  |  | 0.297 |
| No | 0.62 | (0.52, 0.71) |  |
| Yes | 0.57 | (0.54, 0.59) |  |
| Delivery by MTP |  |  | **<0.001** |
| No | 0.53 | (0.50, 0.56) |  |
| Yes | 0.63 | (0.59, 0.66) |  |
| Sex of last child |  |  | 0.655 |
| Girl | 0.56 | (0.53, 0.60) |  |
| Boy | 0.57 | (0.54, 0.61) |  |
| Migration status |  |  | 0.925 |
| Migrant | 0.57 | (0.54, 0.60) |  |
| Non-migrant | 0.57 | (0.53, 0.60) |  |
| Wealth index |  |  | **<0.001** |
| Poor | 0.62 | (0.58, 0.66) |  |
| Middle | 0.58 | (0.53, 0.63) |  |
| Rich | 0.50 | (0.46, 0.53) |  |
| NGO membership |  |  | **<0.001** |
| No | 0.55 | (0.52, 0.57) |  |
| Yes | 0.68 | (0.61, 0.75) |  |
| Place of delivery |  |  | **<0.001** |
| Health facility | 0.65 | (0.62, 0.68) |  |
| Home | 0.43 | (0.39, 0.46) |  |

ANC: Antenatal care, MTP: Medically trained professional, NGO: non-government organization

To find out potential factors associated with pregnancy complications among the urban women in Bangladesh, we selected the best model (Table 4) and conducted multivariate analysis (Table 5), using selected variables that were found to be statistically significant at 5% level in bivariate analysis (Table 3). From Table 2, it can be seen that mean and variance of the count responses (number of pregnancy complications/morbidities) were 0.57 and 0.90 i.e. extra variation exists in the data. It was investigated whether the overdispersion was present or not in pregnancy complications count data used in this study (Table 4). The PR model was first fitted to detect overdispersion by computing the *Dispersion* value using the Pearson chi-square statistic. Table 4 reveals that the *Dispersion* value for the PR model was 1.53 (greater than 1), which clearly depicts the presence of overdispersion and hence, the PR model was found to be overdispersed.

**Table 4:**
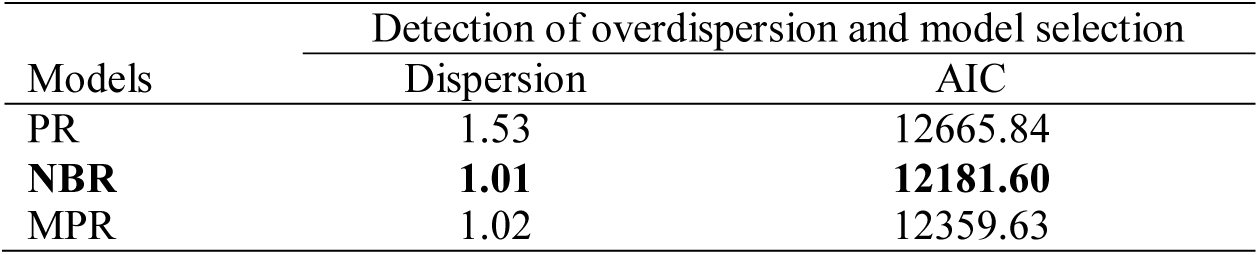
Detection of overdispersion and model selection for analyzing pregnancy complications count response data from Poisson regression (PR), negative binomial regression (NBR) and mixed Poison regression (MPR).

| Models | Detection of overdispersion and model selection |  |
| --- | --- | --- |
|  | Dispersion | AIC |
| PR | 1.53 | 12665.84 |
| **NBR** | **1.01** | **12181.60** |
| MPR | 1.02 | 12359.63 |

**Table 5:**
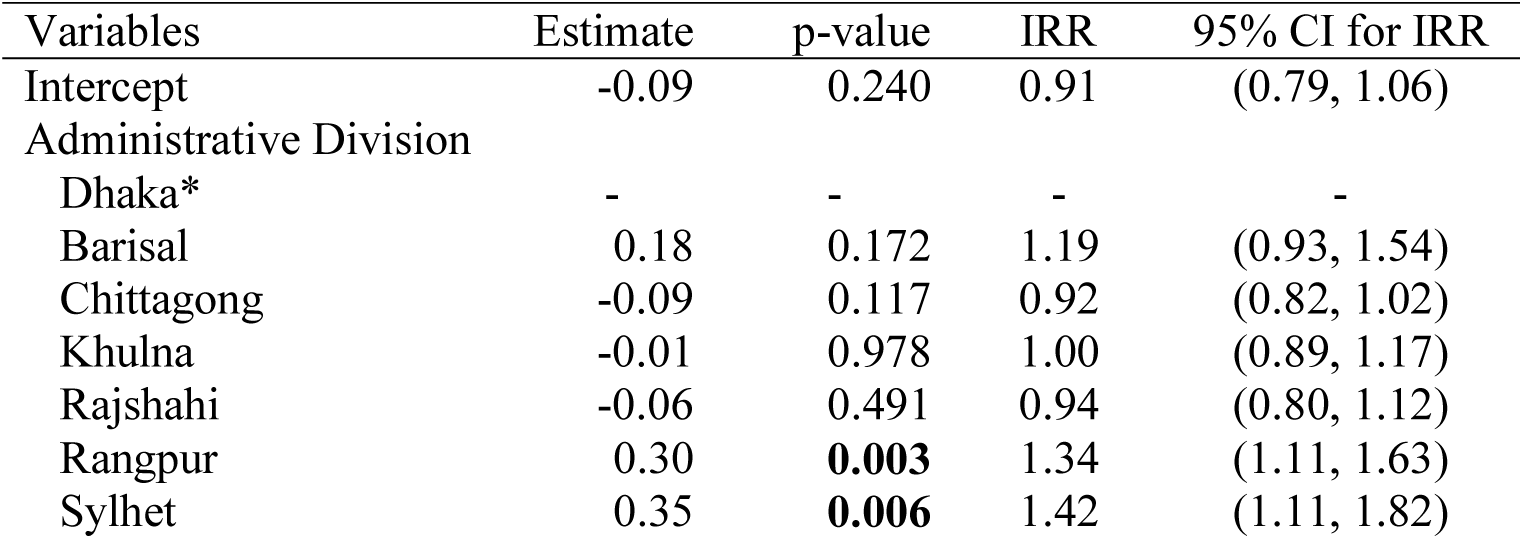
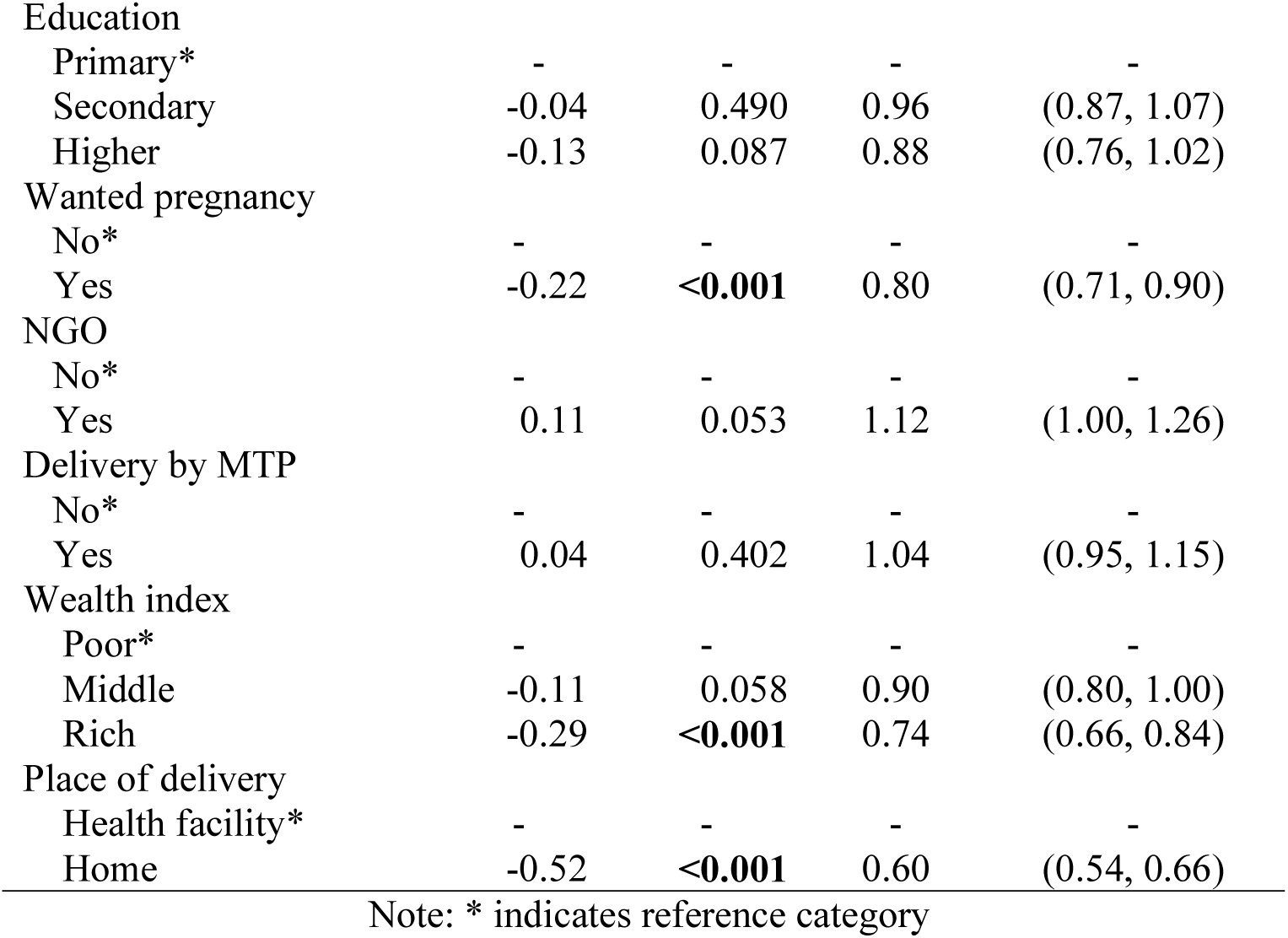
Estimates of parameters, p-values, and incidence rate ratios (IRR) along with 95% confidence interval (CI) for IRR obtained from fitting the negative binomial regression (NBR) to overdispersed maternal morbidity count data of the urban women in Bangladesh.

| Variables | Estimate | p-value | IRR | 95% CI for IRR |
| --- | --- | --- | --- | --- |
| Intercept | -0.09 | 0.240 | 0.91 | (0.79, 1.06) |
| Administrative Division |  |  |  |  |
| Dhaka* | - | - | - | - |
| Barisal | 0.18 | 0.172 | 1.19 | (0.93, 1.54) |
| Chittagong | -0.09 | 0.117 | 0.92 | (0.82, 1.02) |
| Khulna | -0.01 | 0.978 | 1.00 | (0.89, 1.17) |
| Rajshahi | -0.06 | 0.491 | 0.94 | (0.80, 1.12) |
| Rangpur | 0.30 | **0.003** | 1.34 | (1.11, 1.63) |
| Sylhet | 0.35 | **0.006** | 1.42 | (1.11, 1.82) |
| Education |  |  |  |  |
| Primary* | - | - | - | - |
| Secondary | -0.04 | 0.490 | 0.96 | (0.87, 1.07) |
| Higher | -0.13 | 0.087 | 0.88 | (0.76, 1.02) |
| Wanted pregnancy |  |  |  |  |
| No* | - | - | - | - |
| Yes | -0.22 | <0.001 | 0.80 | (0.71, 0.90) |
| NGO |  |  |  |  |
| No* | - | - | - | - |
| Yes | 0.11 | 0.053 | 1.12 | (1.00, 1.26) |
| Delivery by MTP |  |  |  |  |
| No* | - | - | - | - |
| Yes | 0.04 | 0.402 | 1.04 | (0.95, 1.15) |
| Wealth index |  |  |  |  |
| Poor* | - | - | - | - |
| Middle | -0.11 | 0.058 | 0.90 | (0.80, 1.00) |
| Rich | -0.29 | <0.001 | 0.74 | (0.66, 0.84) |
| Place of delivery |  |  |  |  |
| Health facility* | - | - | - | - |
| Home | -0.52 | <0.001 | 0.60 | (0.54, 0.66) |

Note: \* indicates reference category

We then fitted both the NBR and MPR models for further improvement of modelling the overdispersed pregnancy complications count data used in this study. From *Dispersion* and AIC values (Table 4), it was observed that the overdispersion was well captured and modelled by the NBR compared to the PR and MPR models because of its smallest AIC value (12181.60) and approximately 1 (1.01) of the *Dispersion* value. The results obtained from fitting the best model (NBR) were summarized in Table 5.

From Table 5, it is observed that the urban women from Rangpur (*p=*0.003, IRR=1.34, 95% CI: 1.11 to 1.63) and Sylhet (*p=*0.006, IRR=1.42, 95% CI: 1.11 to 1.82) divisional regions were more likely to develop pregnancy complications than women from Dhaka city. Wanted pregnancy (*p*<0.001, IRR=0.80, 95% CI: 0.71 to 0.90) was strongly associated with their average number pregnancy complications/morbidities. As expected, the women who desired the index pregnancy were less likely to experience complications during pregnancy compared to the women who did not. The rich women (*p*<0.001, IRR=0.74, 95% CI: 0.66 to 0.84) were less likely to suffer from pregnancy complications/morbidities than the women who belong to the lower income group.

Place of delivery (*p*<0.001, IRR=0.60, 95% CI: 0.54 to 0.66) was significantly associated with pregnancy complications of the women from urban areas of Bangladesh. However, it was surprising that the women who had delivery at the home experiencing fewer complications than who had at the health facility centre. This may happen because the urban women in Bangladesh visited the health centre when they had already faced pregnancy related complications.

## Discussion and conclusion

Like other developing countries, a significant number of the urban women of Bangladesh experiences major high-risk and life-threating complications/morbidities during their pregnancy and delivery period. This study mainly aims on investigating the prevalence and determinants of maternal morbidities/complications of the urban women during their pregnancy in Bangladesh.

This study findings reveal that 3.3% and 2.7% of the urban women faced life-threating pregnancy and delivery related complications/morbidities, hemorrhage/severe bleeding and fits/convulsion, respectively. Overall, 13.5% of the women experienced at least two complications during their last childbirth. Overdispersion was detected in the count responses of pregnancy complications and therefore, the data were analyzed by fitting the best model, negative binomial regression, because of its minimum AIC value. Whether desired the index pregnancy [11], place of residence or administrative division, place of delivery and wealth index [10] of the urban women were found to be significant risk-factors of maternal morbidity during their pregnancy in Bangladesh.

Women with undesired the index pregnancy, belong to poor/middle income group; and residing in Rangpur/Sylhet regional cities have greater risk of experiencing maternal morbidities or complications during their pregnancy in urban areas of Bangladesh. It is surprising that women who give births at health center are more likely to have complications than their counterparts. This may happen because those urban women in Bangladesh usually visit health centre when they already suffered from pregnancy related complications.

We used the data on maternal morbidity collected in 2013 by the Urban Health Survey as no further survey was conducted yet in Bangladesh, which is the limitation of the study. This study recommends that governmental and nongovernmental organizations, policymakers, and other relevant authorities should focus on the awareness of antenatal health care of women to reducing pregnancy and delivery related complications, and hence maternal and child deaths.

## Data Availability

The Bangladesh Urban Health Survey (BUHS) 2013 https://dataverse.unc.edu/

## Acknowledgements

The authors would like to acknowledge the Urban Health Survey 2013 for providing freely access to their data. The authors also acknowledge the University of Dhaka and University of Grants Commission, Bangladesh for funding this work. We would like to express our gratitude to the Department of Statistics, University of Dhaka, Bangladesh for the technical support during this research project.

## Competing interests

The authors declare that no competing interests exist.

## Funding

This work was funded by the University of Dhaka and University of Grants Commission, Bangladesh.

## Author Contributions

**Conceptualization:** Zakir Hossain, Nilima Afroz, Enamul Kabir

**Data curation:** Zakir Hossain, Nilima Afroz

**Formal analysis**: Zakir Hossain.

**Supervision:** Sabina Sharmin, Sayema Sharmin, Enamul Kabir

**Writing – original draft:** Zakir Hossain, Nilima Afroz

**Writing – review & editing:** Zakir Hossain, Nilima Afroz, Sabina Sharmin, Sayema Sharmin, Enamul Kabir.

## Data availability statement

The secondary data set was used in this study and freely available online at https://dataverse.unc.edu/.

## Notes

### Competing Interest Statement

The authors have declared no competing interest.

### Funding Statement

This work was partially funded by the University of Dhaka and University of Grants Commission, Bangladesh.

## References

1. Bangladesh Urban Health Survey (BUHS) 2013 Final Report. National Institute of Population Research and Training, International Center for Diarrhoeal Disease Research, Bangladesh, Measure Evaluation. Dhaka, Bangladesh and Chapel Hill, North Carolina (USA); 2015. https://www.measureevaluation.org/publications/tr-15-117

2. 2006 Bangladesh Urban Health Survey (BUHS). National Institute of Population Research and Training (NIPORT), MEASURE Evaluation, International Centre for Diarrhoeal Disease Research, Bangladesh (ICDDR,B), and Associates for Community and Population Research (ACPR). Dhaka, Bangladesh and Chapel Hill, NC, USA: NIPORT, MEASURE Evaluation, ICDDR,B, and ACPR; 2008. https://www.measureevaluation.org/resources/publications/tr-08-68/at_download/document

3. Understanding Urban Inequalities in Bangladesh: A prerequisite for achieving Vision 2021. A study based on the results of the 2009 Multiple Indicator Cluster Survey. http://www.indiaenvironmentportal.org.in/files/Urban_paper_lowres.pdf

4. Jolly SP, Rahman M, Afsana K, Yunus FM, Chowdhury AMR. Evaluation of maternal health service indicators in urban slum of Bangladesh. PLoS One. 2016;11(10). https://doi.org/10.1371/journal.pone.0162825

5. Hogan MC, Foreman KJ, Naghavi M, Ahn SY, Wang M, Makela SM. Maternal mortality for 181 countries, 1980-2008: a systematic analysis of progress towards millennium development goal. Lancet. 2010; 375:1609–23. https://doi.org/10.1016/S0140-6736(10)60518-1

6. Lewis G. Beyond the numbers: reviewing maternal deaths and complications to make pregnancy safer. British Medical Bulletin.2003; 67(1):27–37. https://doi.org/10.1093/bmb/ldg009

7. WHO, UNICEF, UNFPA, World Bank Group and the United Nations Population Division. Trends in maternal mortality: 1990 to 2015. https://www.unfpa.org/sites/default/files/pub-pdf/9789241565141_eng.pdf. Accessed on May 15, 2021

8. Sustainable Development Goals. Goal 3: Ensure healthy lives and promote well-being for all at all ages. https://www.un.org/sustainabledevelopment/health/. Accessed on June 20, 2021

9. Say L, Chou D, Gemmill A, Tunçalp Ö, Moller AB, Daniels JD, et al. Global Causes of Maternal Death: A WHO Systematic Analysis. Lancet Global Health. 2014; 2(6):323–33. https://doi.org/10.1016/s2214-109x(14)70227-x

10. Chowdhury ME, Botlero R, Koblinsky M, Saha SK, Dieltiens G, Ronsmans C. Determinants of reduction in maternal mortality in Matlab, Bangladesh: a 30-year cohort study. Lancet. 2007; 370 (9595):1320–1328. https://doi.org/10.1016/s0140-6736(07)61573-6

11. Islam M, Sultana N. Risk factors for pregnancy related complications among urban slum and non-slum women in Bangladesh. BMC Pregnancy Childbirth. 2019; 19(235). https://doi.org/10.1186/s12884-019-2392-6

12. Islam MA, Chowdhury RI, Chakraborty N, Bari W. A multistage model for maternal morbidity during antenatal, delivery and postpartum periods. Statistics in Medicine. 2004; 23:137–158. https://doi.org/10.1002/sim.1594

13. Latif AHMM, Hossain MZ, Islam MA. Model Selection Using Modified Akaike’s Information Criterion: An Application to Maternal Morbidity Data. Austrian Journal of Statistics.2008; 37(2): 175–184. https://doi.org/10.17713/ajs.v37i2.298

14. Chowdhury RI, Islam MA, Gulshan J, Chakraborty N. Delivery complications and healthcare-seeking behaviour: the Bangladesh Demographic Health Survey, 1999–2000. Health and Social Care in the Community. 2007;15(3): 254–264. https://doi.org/10.1111/j.1365-2524.2006.00681.x

15. Chowdhury RI, Islam MA, Chakraborty N, Akhter HH. Determinants of Antenatal Morbidity: A Multivariate Analysis. World Health & Population. 2007; 9(3):9–18. http://doi.org/10.12927/whp.2007.19038

16. Islam MM, Masud MS. Determinants of Frequency and Contents of Antenatal Care Visits in Bangladesh: Assessing the Extent of Compliance with the WHO Recommendations. PloS ONE. 2018; 13(9). https://doi.org/10.1371/journal.pone.0204752.

17. Hossain Z, Maria. Analyzing Overdispersed Antenatal Care Count Data in Bangladesh: Mixed Poisson Regression with Individual-Level Random Effects. Austrian Journal of Statistics. 2021; 50(4):78–90. http://dx.doi.org/10.17713/ajs.v50i4.1163

18. Hilbe JM. Negative Binomial Regression. 2nd ed. Cambridge University Press; 2011.

19. McCullagh P, Nelder JA. Generalized Linear Models. 2nd ed. London: Chapman & Hall; 1989.

20. Dobson AJ, Barnett AG. An Introduction to Generalized Linear Models. 4th ed. CRC press; 2018.

21. Hoef JMV, Boveng PL. Poisson vs. Negative Binomial Regression: How Should We Model Overdispersed Count Data? Ecology. 2007; 88(11):2766–2772. https://www.jstor.org/stable/27651434

22. Ha D. Maximum likelihood estimation using Laplace approximation in Poisson GLMMs. Communications of the Korean Statistical Society. 2009; 16(6):971–978.

23. McCulloch CE, Searle SR, Neuhaus JM (2008). Generalized, Linear, and Mixed Models. 2nd ed. Wiley & Sons, New York; 2008.

24. Mullah MAS, Parveen N, Hossain MZ. Generalized Linear Mixed Models for Longitudinal Data Analysis: An Application to Maternal Morbidity Data. The Dhaka University Journal of Science. 2010; 58(2):219–223.

25. Akaike H. Information theory and an extension of the maximum likelihood principle. In Second International Symposium on Information Theory. Akademia Kiado Budapest. 1973:267–281.

26. Hossain Z, Akter R, Sultana N, Kabir E. Modelling Zero-Truncated Overdispersed Antenatal Health Care Count Data of Women in Bangladesh. PloS ONE. 2020; 15(1):e0227824. https://doi.org/10.1371/journal.pone.0227824.

